# Differences in the genetic architecture of common and rare variants in childhood, persistent and late-diagnosed attention deficit hyperactivity disorder

**DOI:** 10.1101/2021.08.06.21261679

**Authors:** Veera M. Rajagopal, Jinjie Duan, Laura Vilar-Ribó, Jakob Grove, Tetyana Zayats, J. Antoni Ramos-Quiroga, F. Kyle Satterstrom, María Soler Artigas, Jonas Bybjerg-Grauholm, Marie Bækvad-Hansen, Thomas D. Als, Anders Rosengren, Mark J. Daly, Benjamin M. Neale, Merete Nordentoft, Thomas Werge, Ole Mors, David M. Hougaard, Preben B. Mortensen, Marta Ribasés, Anders D. Børglum, Ditte Demontis

**Affiliations:** Department of Biomedicine (Human Genetics) and Centre for Integrative Sequencing, iSEQ, Aarhus University, Aarhus, Denmark; The Lundbeck Foundation Initiative for Integrative Psychiatric Research, iPSYCH, Denmark; Center for Genomics and Personalized Medicine, Aarhus, Denmark; Analytic and Translational Genetics Unit, Massachusetts General Hospital, Boston, MA, 02114, USA; Stanley Center for Psychiatric Research, Broad Institute of MIT and Harvard, Cambridge, MA, 02142, USA; Psychiatric Genetics Unit, Group of Psychiatry, Mental Health and Addiction, Vall d’Hebron Research Institute (VHIR), Universitat Autònoma de Barcelona, Barcelona, Spain; Department for Congenital Disorders, Statens Serum Institut, Copenhagen, Denmark; NCRR - National Centre for Register-Based Research, Business and Social Sciences, Aarhus University, Aarhus, Denmark; Centre for Integrated Register-based Research, CIRRAU, Aarhus University, Aarhus, Denmark; Mental Health Centre Sct. Hans, Capital Region of Denmark, Institute of Biological Psychiatry, Copenhagen University Hospital, Copenhagen, Denmark; Mental Health Centre Copenhagen, Capital Region of Denmark, Copenhagen University Hospital, Copenhagen, Denmark; Psychosis Research Unit, Aarhus University Hospital-Psychiatry, Denmark; Center for Neonatal Screening, Department for Congenital Disorders, Statens Serum Institut, Copenhagen, Denmark; Program in Medical and Population Genetics, Broad Institute of Harvard and MIT, Cambridge, Massachusetts, USA; Institute for Molecular Medicine Finland, University of Helsinki, Helsinki, Finland; Department of Psychiatry, Hospital Universitari Vall d’Hebron, Barcelona, Spain; Biomedical Network Research Centre on Mental Health (CIBERSAM), Instituto de Salud Carlos III, Madrid, Spain; Department of Psychiatry and Forensic Medicine, Universitat Autònoma de Barcelona, Barcelona, Spain; Department of Genetics, Microbiology, and Statistics, Faculty of Biology, Universitat de Barcelona, Barcelona, Spain; BiRC, Bioinformatics Research Centre, Aarhus University, Denmark; PROMENTA, Department of Psychology, University of Oslo, Oslo, Norway

## Abstract

Attention deficit hyperactivity disorder (ADHD) is a neurodevelopmental disorder, with onset in childhood (“childhood ADHD”), and around two thirds of affected individuals will continue to have ADHD symptoms in adulthood (“persistent ADHD”). Age at first diagnosis can vary, and sometimes ADHD is first diagnosed in adulthood (“late-diagnosed ADHD”).

In this study, we analyzed a large Danish population-based case-cohort generated by iPSYCH in order to identify common genetic risk loci and perform in-depth characterization of the polygenic architecture of childhood (N=14,878), persistent (N=1,473) and late-diagnosed ADHD (N=6,961) alongside 38,303 controls. Additionally, the burden of rare protein truncating variants in the three groups were evaluated in whole-exome sequencing data from a subset of the individuals (7,650 ADHD cases and 8,649 controls). We identified genome-wide significant loci associated with childhood ADHD (four loci) and late-diagnosed ADHD (one locus). In analyses of the polygenic architecture, we found higher polygenic score (PGS) of ADHD risk variants in persistent ADHD (mean PGS=0.41) compared to childhood (mean PGS=0.26) and late-diagnosed ADHD (mean PGS=0.27), and we found a significant decreased genetic correlation of late-diagnosed ADHD with inattention (r_g_=0.57) compared to childhood ADHD (r_g_=0.86). These results suggest that a higher ADHD polygenic risk burden is associated with persistence of symptoms, and that a later diagnosis of ADHD could be due in part to genetic factors. Additionally, childhood ADHD demonstrated both a significantly increased genetic overlap with autism compared to late-diagnosed ADHD as well as the highest burden of rare protein-truncating variants in highly constrained genes among ADHD subgroups (compared to controls: beta=0.13, P=2.41×10^−11^). Late-diagnosed ADHD demonstrated significantly larger genetic overlap with depression than childhood ADHD and no increased burden in rare protein-truncating variants (compared to controls: beta=0.06). Overall, our study finds genetic heterogeneity among ADHD subgroups and suggests that genetic factors influence time of first ADHD diagnosis, persistence of ADHD and comorbidity patterns in the sub-groups.

## INTRODUCTION

Attention deficit hyperactivity disorder (ADHD) is a childhood neurodevelopmental disorder characterized by age-inappropriate levels of hyperactivity, impulsivity and inattention. The disorder affects around 5-6% of school-age children, and around 3% of adults^1,2^. It is a complex disorder with both environmental and genetic factors contributing to the risk. Genetics explain a large part of the etiology with an estimated twin heritability of 0.74^3^, and the contribution from common genetic variants is substantial, explaining 22% of the phenotypic variance^4^.

Around two-thirds of children diagnosed with ADHD will continue to have symptoms in adulthood^5^, which is referred to as “persistent ADHD”. Persistent ADHD is associated with more severe outcomes compared to the one-third of individuals that don’t have ADHD as adults (remitters), e.g. increased risk of substance use disorders^6,7^, nicotine dependence^8^ and comorbidity with other psychiatric disorders^9-11^. Several studies have reported a lower heritability for persistent ADHD than for childhood ADHD^12,13^, however these findings have been questioned due to methodological differences of assessment between children and adults^12,14^. It has also been suggested that persistence of symptoms has a genetic risk component specific to persistence rather than base line symptoms^15^ and that a trajectory of persistent symptoms is associated with a high load of common ADHD risk variants^16^.

According to the ICD10 diagnosis criteria, ADHD is a childhood onset disorder, and the behavioral symptoms should be present prior to 7 years of age (prior to 12 years of age according to DSM-5 diagnosis criteria) with a duration of at least six months. However, the disorder is often diagnosed in adolescence and can also be diagnosed in adult life. ADHD symptoms such as hyperactivity and inattention are believed to have continuous distributions in the population, with diagnosed ADHD representing the extreme end, which is supported by genetic findings^4^. The symptoms have an impairing impact on an individual’s life when the accumulation of environmental and genetic risk factors exceeds a threshold. Age at first diagnosis might therefore differ depending on when this threshold is passed. Environmental factors influencing this could be age-related, such as increased educational demands in college or university, resulting in “late-diagnosed ADHD” (i.e. diagnosis in adulthood). Age at first diagnosis might also be influenced by genetic factors affecting symptom heterogeneity and/or severity, and a recent study found individuals with late-diagnosed ADHD to have a burden of common ADHD risk alleles at a level comparable to individuals without ADHD^17^. However, the sample size was very small (N=98 for late-diagnosed individuals), and further investigations is needed in order to elucidate the impact of genetics on age at first diagnosis and determine whether individuals diagnosed with ADHD in adulthood differ genetically from individuals diagnosed as children.

We have previously performed a large genome-wide association study (GWAS) to evaluate the genetic architecture of childhood and persistent ADHD including in total 17,149 ADHD cases and 32,411 controls^18^. The genetic correlation between the two groups was high (r_g_ = 0.81) suggesting that childhood and persistent ADHD to a large extent have the same underlying genetic architecture, however we also noticed that the genetic correlation was significantly different from 1 (P=0.02), suggesting that further dissection of the genetic architecture might reveal genetic differences. Moreover, in that study all adult individuals with ADHD were grouped together, meaning that the persistent group consisted of individuals diagnosed in childhood with persisting symptoms in adulthood and individuals diagnosed as adults (i.e. late-diagnosed ADHD). Further subgrouping of individuals with ADHD depending on age at first diagnosis could therefore reveal unknown information about the genetic architecture underlying the disorder and comorbidities.

In this study we perform in-depth characterization of the polygenic architecture of childhood, persistent and late-diagnosed ADHD in a large Danish population-based cohort of ADHD cases and controls generated by iPSYCH^19^. We observe a higher polygenic burden of ADHD risk variants in persistent ADHD compared to childhood and late-diagnosed ADHD, and find a decreased genetic correlation of late-diagnosed ADHD with ADHD symptoms (inattention and hyperactivity), compared to childhood ADHD. Several significant differences in genetic overlap of ADHD subgroups with other phenotypes are identified, including an increased load of autism risk variants in individuals with childhood compared to late-diagnosed ADHD and a larger genetic overlap of persistent and late-diagnosed ADHD with depression compared to childhood ADHD. Finally, analyses of exome-sequencing data show a significant increased burden of rare protein truncating variants in highly constrained genes in childhood and persistent ADHD compared to controls. This at a rate that that was twice as high as observed for late-diagnosed ADHD, which did not show significant enrichment.

## RESULTS

### Sample characteristics

Individuals with ADHD were identified in the large nation-wide population-based case-cohort established by iPSYCH^19^ consisting of 133,296 genotyped individuals. Individuals with ADHD were identified by information in the Danish Psychiatric Central Research Register^20^ (ICD10 diagnosis code F90.0). ADHD cases were divided into three groups depending on age at first diagnosis: (1) childhood ADHD (N=14,878), defined as individuals diagnosed with ADHD before 18 years of age when the register information was obtained (2016) (2) persistent ADHD (N=1,473), defined as individuals that received an ADHD diagnosis as a child (younger than 18 years of age) and again as adults (older than 18 years of age) and (3) late-diagnosed ADHD (N=6,961), defined as individuals with ADHD that received their first diagnosis as adults (older than 18 years of age). Controls were randomly selected from the same nationwide birth cohort and not diagnosed with ADHD (N=38,303). The sample sizes given above are the number of individuals which remained after quality control (see methods).

The sex distribution was different in the three groups, as females composed 23% of childhood ADHD cases, 36% of persistent ADHD cases and 41% of late-diagnosed cases, and the male/female ratio was significantly different among all three groups (Supplementary Table 1).

Also, comorbidity patterns were different in the three groups. Autism spectrum disorder was very frequent in childhood (23% comorbid) and persistent (18% comorbid) ADHD compared to late-diagnosed ADHD (6.2% comorbid) (Supplementary Table 2). The adolescence/adulthood onset disorders schizophrenia, bipolar disorder and major depressive disorder were more frequent among individuals with persistent and late-diagnosed ADHD. As much as 27% of individuals with late-diagnosed ADHD had comorbid major depressive disorder (Supplementary Table 2).

### Genome-wide association analyses of ADHD subgroups

We conducted a GWAS for each of the three ADHD subgroups against the same control group. The analyses were restricted to a genetically homogenous sample of unrelated individuals with European ancestry and high-quality genetic variants, and corrected for relevant covariates (sex and ancestry covariates).

Four genome-wide significant loci were identified in the GWAS of childhood ADHD located on chromosome 1, 5, 18 and 20 (Table 1, Supplementary Figure 1.A. Supplementary Figure 2. A-D). These consisted of two new ADHD risk loci (on chromosomes 18 and 20) and two known risk loci (on chromosomes 1 and 5), identified in our previous GWAS meta-analysis of ADHD^4^, which included an earlier and smaller iPSYCH sample than analyzed here. One genome-wide significant locus was identified for late-diagnosed ADHD on chromosome 7, located in *FOXP2* (Table 1, Supplementary Figure 1.B. Supplementary Figure 2.E). No genome-wide significant loci were found for persistent ADHD, which was expected due the low number of cases.

**Table 1.**
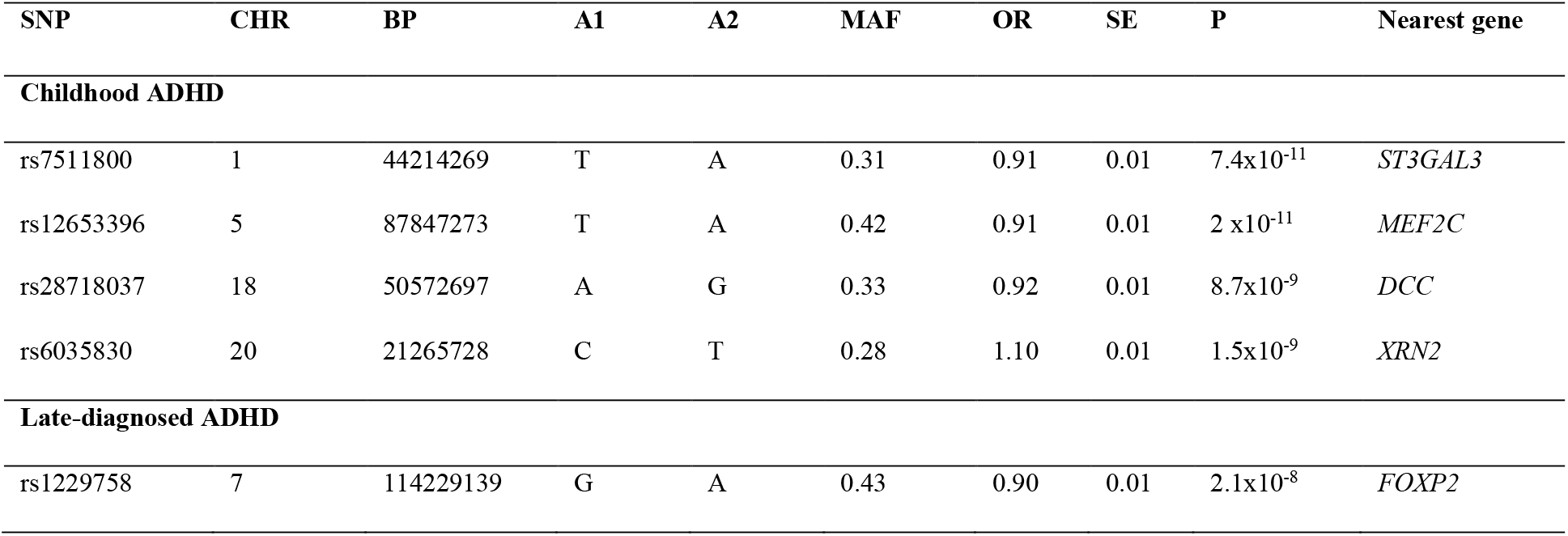
Index variants for the genome-wide significant loci identified in the GWASs of childhood and late-diagnosed ADHD. The variant ID (SNP), chromosome position (CHR), base position in hg19 (BP), effect allele (A1), other allele (A2), minor allele frequency of A1 (MAF), odds ratio (OR), standard error (SE), association P-value from logistic regression (P) and nearest gene is given.

### SNP-heritability and genetic correlations between ADHD subgroups

We estimated the SNP heritability (h^2^_SNP_) using best guess genotypes and GCTA^21^ and found the highest h^2^_SNP_ in the persistent group (h^2^_SNP_ =0.29), followed by late-diagnosed ADHD (h^2^_SNP_ =0.27) and childhood ADHD (h^2^ =0.24) (Supplementary Table 3). None of the estimates were significantly different (Supplementary Table 3).

Pair-wise genetic correlations between ADHD subgroups were estimated using bivariate GREML implemented in GCTA and non-overlapping controls (see methods). A high genetic correlation was found between childhood and persistent ADHD (r_g_=0.82, SE=0.08), as well as between persistent and late-diagnosed ADHD (r_g_=0.77, SE=0.08), while the genetic correlation between childhood ADHD and late-diagnosed ADHD was moderate (r_g_=0.64, SE=0.03), but not significantly lower than the other two correlations (Supplementary Table 4).

### ADHD polygenic risk load in childhood, persistent and late-diagnosed ADHD

The polygenic risk load of common variants associated with ADHD risk in the three ADHD subgroups was evaluated by polygenic score (PGS) analyses. ADHD-PGS was generated using a 5-fold cross-validation approach (see online methods) and regressed on the three ADHD subgroups and controls while including relevant covariates in a multiple regression. All groups demonstrated a highly significant increased ADHD-PGS load compared to controls (Supplementary Table 5). The highest mean ADHD-PGS was found for persistent ADHD (mean=0.41, SE=0.95), followed by late-diagnosed ADHD (mean=0.27; SE=0.98) and then childhood ADHD (mean=0.26, SE=0.96) (Supplementary Table 5). ADHD-PGS load in persistent ADHD was significantly higher childhood ADHD (P=3.0×10^−4^) and nominally significantly higher than late-diagnosed ADHD (P=0.02).

In order to replicate the findings, we performed PGS analysis in a Spanish sample consisting of 453 individuals with childhood ADHD, 270 with persistent ADHD, and 889 with late-diagnosed ADHD and 3,440 controls. We did not replicate the findings observed in the iPSYCH data, however the differences were non-significant with trends in the opposite direction when comparing to controls (ADHD-PGS childhood ADHD: beta=0.27, SE=0.05; persistent ADHD: beta=0.21, SE=0.06; late-diagnosed ADHD: beta=0.19, SE=0.04).

### Genetic overlap with ADHD symptoms in the general population

Genetic overlap with ADHD symptoms in the general population was estimated, using results from GWASs of ADHD subgroups and results from GWAS meta-analyses of measures of inattention and hyperactivity (N=43,117) in the general population (Zayates et al. unpublished data). Inattention and hyperactivity were highly correlated with both childhood ADHD (r_g_inattanetion_=0.86, SE=0.08; r_g_hyperactivity_=0.95, SE=0.08) and persistent ADHD (r_g_inattention_=0.87, SE=0.14; r_g_hyperactivity_=∼1, SE=0.15) (Supplementary Table 6), but showed a considerably lower correlation with late-diagnosed ADHD (r_g_inattention_ = 0.57, SE =0.08; r_g_hyperactivity_=0.59, SE=0.07). The genetic correlations of ADHD symptoms with childhood and persistent ADHD were not different from one, while the genetic correlations with late-diagnosed ADHD were significantly lower than one (P_diff_1_inattention_=7.66×10^−8^; P_diff_1_hyperactivity_=4.71×10^−9^; Supplementary Table 6).

We also carried out PGS analyses to test for enrichment in the three ADHD subgroups of variants associated with inattention and hyperactivity. We identified a nominally significantly lower PGS for hyperactivity in late-diagnosed ADHD compared to childhood ADHD (P=0.04; Supplementary Table 7).

### Genetic overlap with psychiatric disorders and other phenotypes

The observed differences in comorbidity patterns with other psychiatric disorders could reflect age differences among the groups, but they could also be influenced by differences in the genetic architecture. In order to evaluate whether genetics influence the different comorbidity patterns, we performed genetic correlation and PGS analyses for major psychiatric disorders (schizophrenia^22^, bipolar disorder^23^, major depressive disorder^24^, autism spectrum disorder^25^, anorexia^26^, obsessive compulsive disorder^27^, cannabis use disorder^28^ and alcohol use disorder^29^). We found positive genetic correlations of ADHD subgroups with autism, schizophrenia, bipolar disorder, major depressive disorder, alcohol use disorder and cannabis use disorder and negative genetic correlations with obsessive compulsive disorder and anorexia (Figure 1, supplementary Table 8), in line with previous findings (e.g^4^). We identified a significantly higher genetic correlation of childhood ADHD with autism (r_g_=0.48, SE=0.05) compared to late-diagnosed ADHD (r_g_=0.27, SE=0.06) (Jack knife comparison P-value = 4.9×10^−4^), and a significantly (P=8.7×10^−7^) higher genetic correlation of depression and alcohol use disorder with late-diagnosed ADHD (r_g_depression_=0.69, SE=0.04; r_g_alcohol_use_disorder_=0.45, SE=0.04) compared to childhood ADHD (P_diff_depression_=8.7×10^−7^; P_diff_alcohol_use disorder_=3.8×10^−5^) (Figure 1, Supplementary 9 Table 8). The PGS results demonstrated the same pattern, with a significantly increased autism-PGS in childhood ADHD compared to late-diagnosed ADHD (P=7.87×10^−7^), higher PGS in persistent and late-diagnosed ADHD compared to childhood ADHD for depression (P_childhood_vs_persistent_ < 0.1×10^−15^; P_childhood_vs_late-diagnosed_=1.95×10^−9^) and cannabis use disorder (P_childhood_vs_persistent_=9.02×10^−5^; P_childhood_vs_late-diagnosed_=1.42×10^−5^) and significantly increased PGS in late-diagnosed ADHD compared to childhood ADHD for schizophrenia (P=2.22×10^−4^) and bipolar disorder (P=9.34×10^−7^) (Figure 1, Supplementary Table 9). Additionally, we performed genetic correlation and PGS analyses for phenotypes representing domains which previously^4^ demonstrated high genetic correlations with ADHD: cognition (educational years^30^), overweight (BMI^31^), reproduction (age at first birth^32^), mortality (maternal age of death^33^) and sleep (insomnia^34^). We identified stronger negative genetic correlations of late-diagnosed ADHD compared to childhood ADHD for educational years (P_difference_=1.7×10^−5^; r_g_late-diagnosed_=-0.61, SE=0.03; r_g_childhood_=-0.46, SE=0.03), increased age at first birth (P_difference_=8.9×10^−5^; r_g_late-diagnosed_=-0.73, SE=0.04; r_g_childhood_=-0.54, SE=0.04) and increased age at mother’s death (P_difference_=2.6×10^−4^ r_g_late-diagnosed_=-0.79, SE=0.10; r_g_childhood_=-0.48, SE=0.08) (Figure 1, Supplementary Table 8). Furthermore, we identified a significantly less negative PGS in childhood ADHD compared to persistent and late-diagnosed ADHD for number of educational years (P_childhood_vs_persistent_=8.02×10^−8^; P_childhood_vs_late-diagnosed_=4.35×10^−14^) and a less negative PGS for age at first birth for childhood ADHD compared to late-diagnosed ADHD (Figure 2, Supplementary Table 9).

**Figure 1.**
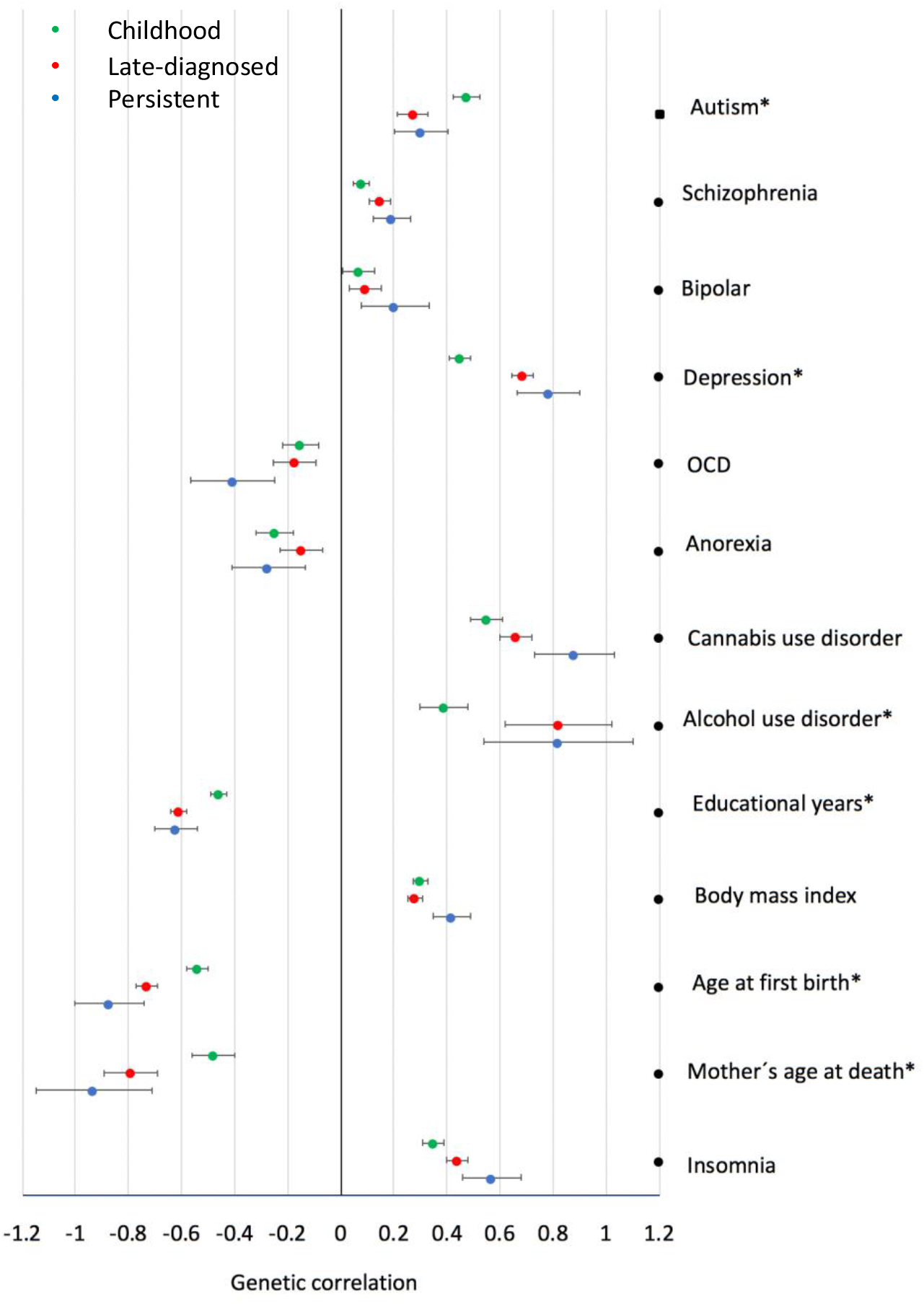
Genetic correlation estimated using LD score regression of childhood, persistent and late-diagnosed ADHD with major psychiatric disorders and phenotypes representing domains with high genetic correlation with ADHD. Horizontal bares represent standard errors. *indicates significant difference in the genetic correlation observed for childhood ADHD when compared to late-diagnosed ADHD (P<0.0013).

**Figure 2.**
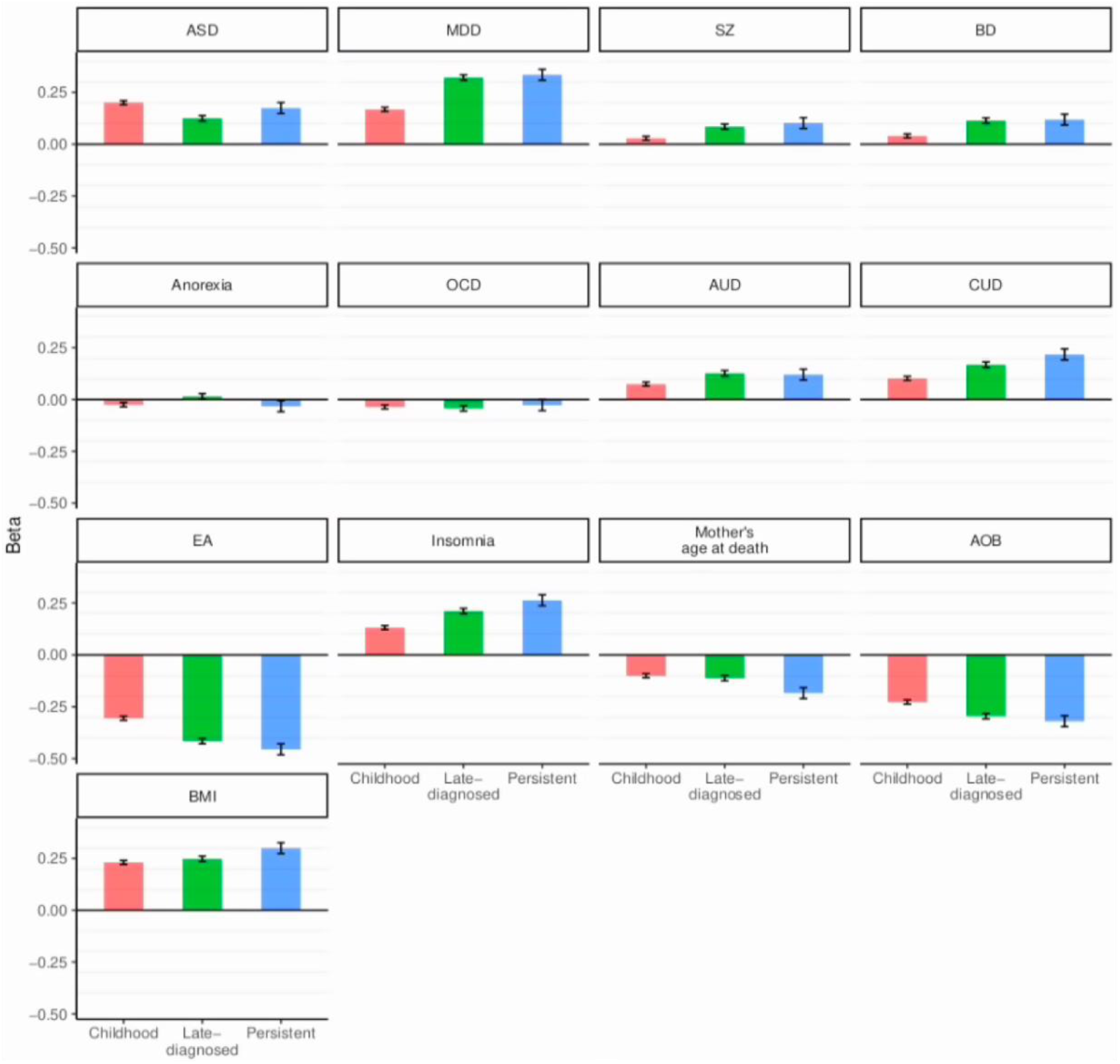
Results from PGS analysis demonstrating the association of PGS with childhood, persistent and late-diagnosed ADHD. PGS for psychiatric disorders: autism spectrum disorder (ASD), depression (MDD), schizophrenia (SZ), bipolar disorder (BD), anorexia, obsessive compulsive disorder (OCD), alcohol use disorder (AUD), cannabis use disorder (CUD). PGS for five phenotypes representing domains highly correlated with ADHD: educational attainment (EA), insomnia, mother’s age at death and age of first birth (AOF). On the y-axis is the beta from multi-nominal regression against controls; vertical bares represent standard errors (See also Supplementary Table 10). For significance between groups see Supplementary Table 9.

Except for autism and OCD, the highest PGS was observed for persistent ADHD (Supplementary Table 10); however, due to the smaller sample size of this group compared to the other two groups, we had limited power to detect differences in PGS load for this group compared to the other two groups.

### Burden of rare deleterious variants in childhood, persistent and late-diagnosed ADHD

We have recently demonstrated an enrichment of rare deleterious variants in highly constrained genes (genes intolerant to loss of function mutations) in ADHD cases compared to controls^35^. We wanted to explore this further by evaluating the load of rare protein truncating variants (rPTVs) in the three ADHD subgroups. For this we used whole-exome sequencing data available for a subset of the iPSYCH cohort (childhood ADHD, N=4,987; persistent ADHD, N=748; late-diagnosed ADHD, N=1,915; controls, N=8,649). The burden of rPTVs and rare synonymous variants (rSYN) in the three ADHD subgroups was tested in three gene sets: (1) “highly constrained genes”, genes being evolutionary intolerant to loss of function mutations with pLI > 0.9^36^ (3,488 genes), (2) “*de novo* constrained genes”, the subset of highly constrained genes that overlap another gene-set of 285 genes found to be enriched with de novo mutations in individuals with neurodevelopmental disorders^37^ (241 overlapping genes) and (3) “low constrained genes”, genes being tolerant to loss of function mutations with a pLI < 0.1 (9,662 genes). The load of rPTVs was comparable for childhood and persistent ADHD in both highly constrained genes (childhood ADHD beta=0.13, SE=0.02; persistent ADHD beta=0.12, SE=0.04) and *de novo* highly constrained genes (childhood ADHD beta=0.1, SE=0.02; persistent ADHD beta=0.09, SE=0.04), but lower in late-diagnosed ADHD (highly constrained: beta=0.06, SE=0.03; *de novo* highly constrained: beta=0.03, SE=0.03) (Supplementary Table 11, Figure 3). We only identified significant enrichment in rPTVs in childhood and persistent ADHD in highly constrained genes (P_childhood_ADHD_=2.41xa10^−11^, P_persistent_ADHD_ = 1.90×10^−3^). No pair-wise comparisons of rPTV load were significant, but a tendency towards a higher burden of rPTVs in childhood compared to late-diagnosed ADHD was observed (P=0.096). For comparison we did not find any enrichment of rSYNs in either highly constrained or *de novo* constrained genes or an enrichment in of rPTVs in low constrained genes (Figure 3, Supplementary Tables 11).

**Figure 3.**
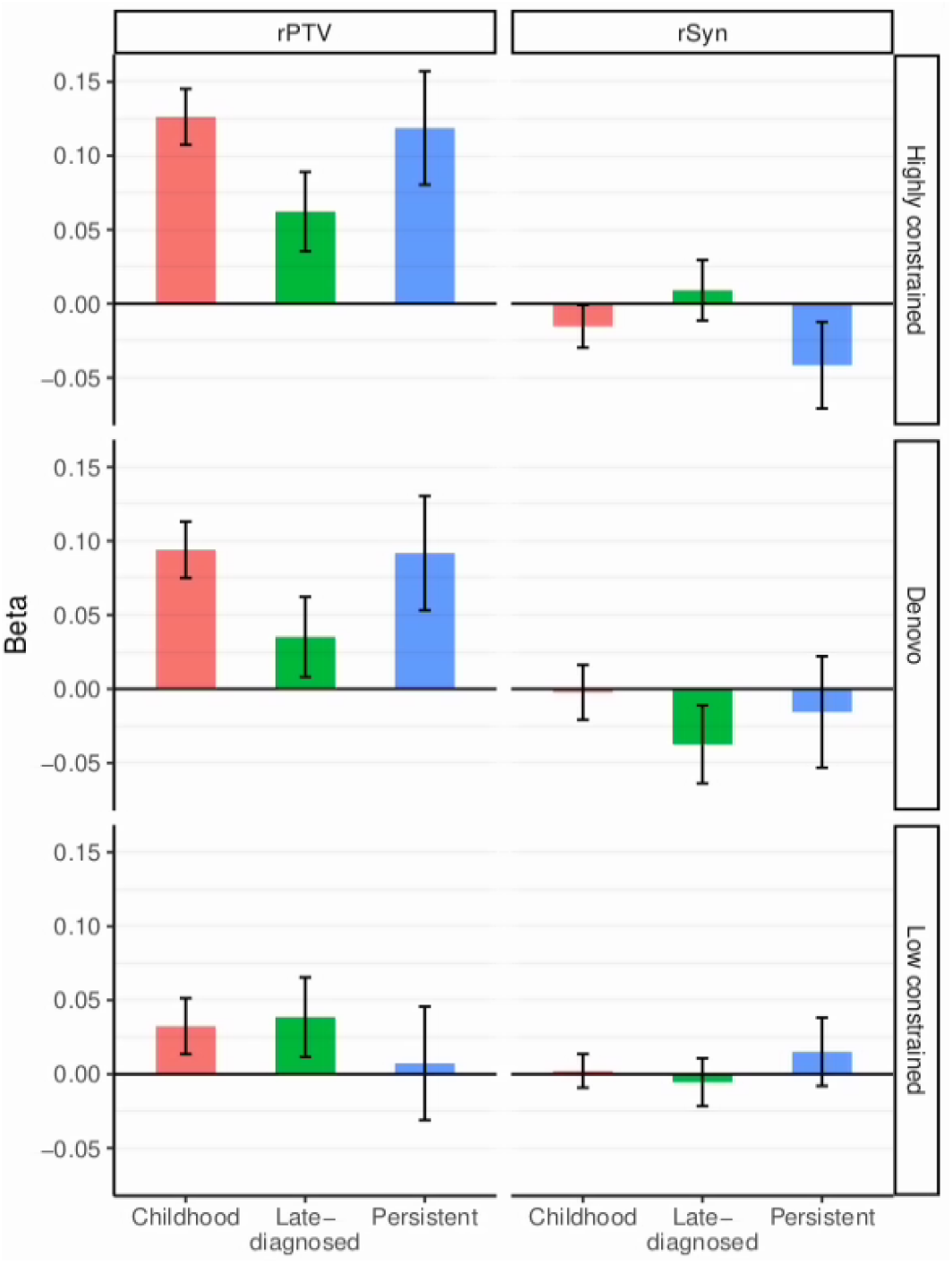
The load of rare protein truncating variants (rPTVs) and rare synonymous (rSyn) in three gene-sets (1) “Highly constrained”, genes intolerant to loss of function mutations (pLI > 0.9), (2) “Denovo”, highly constrained genes which in another study has been found to be enriched with *de novo* mutations in individuals with neurodevelopmental disorders and (3) “Low constrained”, genes tolerant to loss of function mutations (pLI < 0.1). Y-axis represents the beta from multiple logistic regression with comparison to controls, vertical lines represent the standard error. For pair-wise comparisons of ADHD subgroups see Supplementary Table 11.

## DISCUSSION

In this study we identified differences in the genetic architecture of childhood, persistent and late-diagnosed ADHD based on unique data from a large population-based large Danish cohort. We identified four genome-wide significant loci associated with childhood ADHD, two of them novel ADHD risk loci located on chromosome 18 and 20. The chromosome 18 index variant is located *DCC*, a gene recently linked to the general liability to psychiatric disorders in a large cross-disorder study by the Psychiatric Genomics Consortium^38^, and thus does not seem specific to ADHD. The chromosome 20 locus is intergenic, and the index variant has previously shown genome-wide significant association with weight-related phenotypes^33^. These are the first loci associated with childhood ADHD, as our previous meta-analysis of childhood ADHD^18^ did not reveal significant loci. Additionally, we identified a genome-wide significant locus associated with late-diagnosed ADHD in *FOXP2*. When this locus was first reported as a risk locus for ADHD^4^, it received a lot of attention due to the role of *FOXP2* in cognition, language and speech development^39-41^, and recently we also found *FOXP2* to be a risk gene for cannabis use disorder^28^. The results reported here suggest that the association of *FOXP2* with ADHD is driven to a larger extent by late-diagnosed ADHD (OR=1.11) than childhood ADHD (OR=1.05).

When assessing the polygenic architecture, we identified the highest SNP heritability for persistent ADHD (Supplementary Table 3). In concordance with this we also observed the highest polygenic risk load for ADHD and ADHD symptoms (inattention and hyperactivity) in individuals with persistent ADHD, which was significantly higher than observed for childhood ADHD (Supplementary Table 5 and 6). This observation is consistent with the hypothesis that individuals with a higher genetic risk load for ADHD also are those that will continue to have ADHD symptoms as adults. This is also supported by a previous study reporting that ADHD-PGS is associated with persistence of ADHD symptoms across childhood and adolescence in the general population^16^, and a recent study reporting higher ADHD-PGS in persistent ADHD compared to late-diagnosed ADHD (however not significantly different)^17^. We were not able to replicate our findings in a Spanish cohort. Lack of replication could be due to replication cohort size or differences in the way cases have been selected. The iPSYCH cohort is population-based and thus reflects the genetic architecture across cases with ADHD in the entire Danish population, whereas the Spanish cohort is a smaller clinical dataset that might be influenced by unknown ascertainment biases.

The genetic correlation of childhood ADHD with persistent ADHD was high (r_g_=0.82) and at the same level as reported previously (r_g_=0.81)^18^, while the genetic correlation of childhood ADHD with late-diagnosed ADHD was considerably lower (r_g_=0.65). This suggests some differences in the polygenic architecture of childhood and late-diagnosed ADHD. A part of this could be due to a lower load of variants associated with hyperactivity and inattention in individuals with late-diagnosed ADHD, since we observed a higher genetic correlation of ADHD symptoms with childhood and persistent ADHD compared to late-diagnosed ADHD. However, only significant for inattention. The same pattern was observed in the PGS analyses where we found a higher PGS for inattention in childhood and persistent ADHD than in late-diagnosed ADHD, however pair-wise comparisons only revealed nominally significant higher PGS for hyperactivity in childhood ADHD compared to late-diagnosed ADHD. Our results therefore suggest that a part of the explanation for a later diagnosis of ADHD could be genetic, so late-diagnosed individuals genetically are less predisposed to be inattentive and hyperactive and thus their ADHD could be unnoticed until later in life.

The comorbidity pattern in the three groups differed, with a higher comorbidity of autism spectrum disorder among childhood (23%) and persistent ADHD (18%) compared to late-diagnosed ADHD (6.2%), in line with previous reports concerning comorbid autism among children with ADHD^42^. We found a significant genetic overlap between ADHD and autism as previously reported^25,43^, and the observed comorbidity patterns were reflected in the genetic analyses where we found a significantly higher genetic correlation of autism with childhood ADHD compared to the genetic correlation with late-diagnosed ADHD, as well as higher PGS-autism in childhood ADHD compared to late-diagnosed ADHD. Therefore, childhood ADHD seems genetically more related to autism than late-diagnosed ADHD is related to autism.

The comorbidities of psychiatric disorders with onset in adolescence/adulthood were higher among persistent and late-diagnosed ADHD (Supplementary Table 2). Part of this result is likely due to the age difference, since many individuals in the childhood group are too young to develop these disorders. However, our results suggest that age alone cannot explain the different comorbidity patterns. Genetics might play a role, as we in general observe a higher genetic correlation or PGS for several of the disorders (schizophrenia, bipolar disorder, alcohol use disorder, cannabis use disorder and depression) in persistent and late-diagnosed ADHD compared to childhood ADHD (Figure 1 and 2, Supplementary Table 8 and 9). Depression was particularly striking, where analyses revealed large differences with a significantly higher PGS in individuals with persistent and late-diagnosed ADHD compared to childhood ADHD. The high comorbidity of depression among adults with ADHD is well known, but the causes are not. It has been suggested that ADHD in itself could be a risk factor^44,45^, but genetics is also considered a risk factor due to the previously reported high genetic correlation of ADHD and depression^4^. Our results suggest genetic heterogeneity among ADHD cases where individuals with persistent and late-diagnosed ADHD are at higher risk for getting comorbid depression due to the underlying genetic architecture of the disorder in these groups.

In analyses of five selected phenotypes representing domains highly genetically correlated with ADHD^4^ we observed the highest genetic correlations and highest PGS load in persistent ADHD, followed closely by late-diagnosed ADHD and lowest in childhood ADHD (except for BMI), suggesting a similar polygenic architecture of persistent and late-diagnosed ADHD for these phenotypes (Figure 1 and 2). These results also support the idea that the negative outcomes associated with persistent ADHD such as decreased school performance^46^ and sleep problems^47^, are influenced by genetics to a greater extent than in childhood ADHD.

We found a significantly increased burden of rPTVs in persistent and childhood ADHD compared to controls in highly constrained genes, but no increased burden in late-diagnosed ADHD. Additionally, there was a tendency towards a significantly higher burden of rPTVs in *de novo* highly constrained genes in childhood ADHD compared to late-diagnosed ADHD. These findings suggest that when considering rPTVs, which are variants expected to have higher impact on the disorder than common variants, the genome of individuals with late-diagnosed ADHD is less burdened. When considering both common and rare variants, the emerging picture suggests that childhood ADHD is genetically more similar to autism (high genetic correlation with autism and increased rPTV burden), whereas late-diagnosed ADHD genetically is more similar to depression (high genetic correlation with depression and no significant increase in rPTVs in highly constrained genes compared to controls; Supplementary Table 11).

In summary, our results are population-based and thus reflect the genetic architecture of ADHD and comorbidity patterns across ADHD subgroups in the Danish population. Persistent ADHD demonstrated the highest load of ADHD risk variants, while late-diagnosed ADHD was less enriched for variants associated with hyperactivity and inattention and did not, unlike childhood and late-diagnosed ADHD, demonstrate an increased burden of rPTVs compared to controls. This suggests that genetics in part might explain why some individuals are diagnosed late as adults. The comorbidity pattern for other psychiatric disorders differed among the groups, and our results suggest that the higher comorbidity of adult onset psychiatric disorders among individuals with persistent and late-diagnosed ADHD is not only due to those individuals being older, but also due to a higher genetic risk. Conversely, childhood ADHD demonstrated a higher genetic overlap with autism and higher burden of rPTVs in highly constrained genes than the other two groups. Overall, our study finds genetic heterogeneity among ADHD subgroups and suggests that genetic factors influence time of first ADHD diagnosis, persistence of ADHD into adulthood and comorbidity patterns.

## Methods

### Sample characteristics

Individuals included in the study were identified in a large nationwide population-based case-cohort established by iPSYCH^19^ in total comprising 133,296 genotyped individuals, among which 91,378 are cases diagnosed with at least one of six mental disorders (i.e. schizophrenia, bipolar disorder, major depressive disorder, autism spectrum disorder, ADHD, anorexia) and the remaining population-based controls. Samples were selected from a baseline birth cohort comprising all singletons born in Denmark between May 1, 1981, and December 31, 2008, who were residents in Denmark on their first birthday and who have a known mother (N = 1,472,762). ADHD cases were diagnosed by psychiatrists at in- or out-patient clinics according to the ICD10 criteria (F90.0 diagnosis code) identified using the Danish Psychiatric Central Research Register^20^. Diagnoses were given in 2016 or earlier for individuals at least 1 year old. Cases were divided into three groups depending on age at first diagnosis: (1) childhood ADHD, defined as cases diagnosed with ADHD and were less than 18 years of age in 2016 (N=15,338 before QC), (2) Persistent ADHD defined as cases having received an ADHD diagnosis as child (less than 18 years of age) and again as adults (after 18 years of age) (N=1,709 before QC) and (3) late diagnosed ADHD, defined as individuals having received an ADHD diagnosis as adults (after 18 years of age) (N=7,815 before QC). Controls were randomly selected from the same nationwide birth cohort and not diagnosed with ADHD (N=45,398 before QC). The study was approved by the Danish Data Protection Agency and the Scientific Ethics Committee in Denmark.

Testing for difference in the female/male ratio between ADHD subgroups was done using a chi-square test. Information about comorbidity for other major psychiatric disorders was obtained from the Danish Psychiatric Central Research Register^20^: autism spectrum disorder (ICD10 diagnosis code F84), schizophrenia (ICD10 diagnosis code F20), bipolar disorder (ICD10 diagnosis codes F30-F31), major depressive disorder (ICD10 diagnosis codes F32-F33), obsessive compulsive disorder (ICD10 diagnosis code F42), anorexia (ICD10 diagnosis codes F50), alcohol use disorder (ICD10 diagnosis code F10.1-F10.9), cannabis use disorder (ICD10 diagnosis code F12.1-F12.9).

### Genotyping and QC

The study subjects identified in the registers were linked to their biological sample (dried blood spots) stored in the Danish Newborn Screening Biobank^48^, by the use of the personal identification number^49^ assigned to all individuals with residence in Denmark. DNA was extracted from the dried blood spots and whole genome amplified in triplicates as described previously^50,51^. Genotyping of the iPSYCH samples were done in two rounds. In round one (iPSYCH1), 79,492 individuals were genotyped using Illumina’s Beadarrays (PsychChip; Illumina, CA, San Diego, USA) according to the manufacturer’s protocols. In round two (iPSYCYH2) 53,804 individuals were genotyped using Illuminas Global Screening array. iPSYCH genotypes were a result of merging call sets from two different calling algorithms, GenCall^52^ and Birdseed^53^ which were used to call genotypes with minor allele frequency (MAF) > 0.01. iPSYCH2 genotypes were called using GenTrain V3.

Following genotyping, all data processing, quality control, and downstream analyses were performed at our secure server at Aarhus University (GenomeDK (http://genome.au.dk)).

Stringent quality control was applied to the data to the full iPSYCH sample. Only individuals with high call rate (> 0.95) were included, and only genotypes with high call rate (>0.98), no strong deviation from Hardy-Weinberg equilibrium (*P* >1×10^−6^ in controls or *P* >1×10^−10^ in cases) and low heterozygosity rates (| F_het_ | <0.2) were included. Genotypes were phased and imputed using the Haplotype Reference Consortium^54^ data as the imputation reference panel, while pre-phasing was done using Eagle v2.3.5^55^ and imputation using Minimac3^56^.

Relatedness and population stratification were evaluated for ADHD cases and controls using merged data from iPSYCH1 and iPSYCH2 and a set of high quality markers (best guess genotypes with MAF >0.05, HWE P >1×10^−9^, SNP call rate >0.99, imputation info score (INFO) > 0.9), which were pruned for linkage disequilibrium (LD) (r^2^ <0.1) resulting in a set of 37,986 pruned variants (markers located in long-range LD regions defined by Price et al.^57^ were excluded). Genetic relatedness was estimated using PLINK v1.9^58,59^ to identify first and second-degree relatives 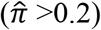, and one individual was excluded from each related pair (cases preferred kept over controls). Genetic outliers were identified for exclusion based on principal component analyses (PCA) using EIGENSOFT^60,61^. After the first PCA, the PCs from a set of individuals born in Denmark for three generations were used as a reference for generating an ellipsoid based on information from the first 6 PCs and their standard deviations (8 SDs were used). Those who fell outside this ellipsoid were removed. Then the PCA was then repeated and the new PCs were used as covariates to correct for any remaining population substructure in the subsequent analyses. After quality control, the number of included individuals is: (1) childhood ADHD; N=14,878 (2) persistent ADHD; N=1,473 (3) late diagnosed ADHD; N=7,188. After QC the control group contained 38,3030 individuals.

### GWAS

We performed GWASs for each ADHD subgroup (childhood N=14,878 cases, persistent N=1,473 cases, late-diagnosed N=7,188 cases) against a common set of controls (N=38,3030). We used merged iPSYCH1 and iPSYCH2 high quality best guess genotypes (MAF>0.01, INFO>0.80; N=5,826,893 variants) and logistic regression and corrected for population structure using 10 PCs from PCA and a covariate for genotyping round (iPSYCH1 or iPSYCH2). The analyses were performed using PLINK1.9^58^.

### SNP-heritability and genetic correlation between ADHD subgroups

SNP-heritability for each ADHD subgroup was estimated using a genotype relatedness matrix only including high quality variants present in both iPSYCH1 and iPSYCH2 (N=5,826,893variants). The SNP heritability was estimated for each group against the same controls (N=38,3030) using univariate GREML implemented in GCTA^21^ and a population prevalence of childhood ADHD of 0.05, of persistent ADHD and of 0.03 and late-diagnosed ADHD of 0.03. In order to test for difference in SNP-heritability estimates among groups, we also derived estimated using non-shared controls (control numbers: childhood controls: 24,443, persistent controls 2,289, late-diagnosed controls 11,571). Testing for difference in SNP-heritability was done using a z-test.

Genetic correlations between ADHD subgroups were calculated using bivariate GREML analysis in GCTA and non-shared controls.

### PGS analyses of ADHD and other phenotypes

The polygenic scores for ADHD were generated using a 5-fold cross-validation approach, similar to what we did previously^4^. In short, the sample was split into five groups, aiming for approximately equal numbers of ADHD cases within each group. We then conducted five leave-one-out analyses, with each leave-one-out analysis using four out of five groups as training datasets for estimation of effect sizes with respect to ADHD risk. These effect sizes were then used to estimate PGS for the remaining target group. Indels and variants in the extended MHC region (chromosome 6: 25-34 Mb) were also removed. Only independent variants were used to generate the score, and clumping of the training data was done on the summary statistics employing PLINK and the flags -clump-p1 1, - clump-p2 1, -clump-r2 0.1 and -clump-kb 500. PGS were estimated for each individual in the target sample using a range of P-value thresholds in the training data (5×10^−8^, 1×10^−6^, 1×10^−4^, 1×10^−3^, 0.01, 0.05, 0.1, 0.2, 0.5, 1.0), multiplying the natural log of the odds ratio of each variant by the allele-count of each variant. Whole genome PGS were obtained by summing values over variants for each individual. For each of the five groups of target samples PGS were standardized (subtracting the mean and dividing by the standard deviation).

PGS for ADHD symptoms and 13 other phenotypes (schizophrenia, autism, bipolar disorder, alcohol use disorder, cannabis use disorder, obsessive compulsive disorder, anorexia, depression, educational years, mother’s age at death, body mass index, age at first birth and insomnia) were generated using summary statistics from large GWASs of the phenotypes (see Supplementary Table 9 for references) and the approach described above (without 5-fold cross validation). The data on ADHD symptoms (inattention and hyperactivity/impulsivity) come from a genome-wide association meta-analysis on up to 43,117 children and adolescents (Zayats et al. unpublished data). In the PGS analyses P-value thresholds in the training GWASs that captured most variance (estimated by Nagelkerke’s *R*^*2*^) in the target data when pooling all ADHD cases together were used as threshold for analyses of PGS load in the subgroups (threshold information in Supplementary Table 9).

We tested for difference in PGS load among ADHD subgroups using multi-nominal regression with ADHD coded as four factors: controls, childhood, adulthood, and persistent and we included covariates to correct for genotyping round and 10 ancestry principal components. Correction for multiple testing was done separately for the following three analyses: (1) PGS-ADHD load among subgroups correcting for three pair-wise comparisons, (2) PGS load for ADHD symptoms (inattention and hyperactivity) correcting for six pair-wise comparisons and (3) PGS load for 13 other phenotypes correcting for 39 pair-wise comparisons.

PGS analysis in the Spanish cohort consisting of 453 individuals with childhood ADHD, 270 with persistent ADHD, 889 with late-diagnosed ADHD and 3,440 controls was done using the same approach as above. A detailed description of ADHD diagnosis procedures for these samples can be found in Rovira et al.^18^. Exclusion criteria for patients were: IQ<70, having pervasive developmental disorders, schizophrenia or other psychotic disorders, adoption, sexual or physical abuse, birth weight<1.5 kg and any significant neurological or systemic disease that might explain ADHD symptoms. Comorbid oppositional defiant disorder, conduct disorder, depression and anxiety disorders were allowed unless determined to be the primary cause of ADHD symptomatology.

### Genetic correlations with ADHD symptoms and other phenotypes

Genetic correlations of the three ADHD subgroups with ADHD symptoms and the 13 phenotypes listed above were calculated using summary statistics from GWASs and LD score regression^62^. The intercept was restricted to 1, when calculating rg with ADHD symptoms as no sample overlap and population stratification was present.

Statistical difference between two r_g_ estimates was calculated using the block jackknife method implemented in the LD score regression software^62,63^. The genome was divided in 200 blocks and jackknife delete values were calculated by excluding one block at a time. The jackknife deleted values were then used to calculate corresponding jackknife pseudo values. Based on the mean and variance of the jackknife pseudovalues, Z-score and corresponding P-values were computed, testing the null hypothesis that the difference between the genetic correlations is equal to zero. Test for difference in the genetic correlation from one, was done using a z-test.

Correction for multiple pair-wise comparisons, was done separately for the following two evaluations: (1) difference in r_g_ of ADHD subgroups with ADHD symptoms correcting for six pair-wise comparisons and (2) r_g_ difference of ADHD subgroups with 13 other phenotypes correcting for 39 pair-wise comparisons.

### Burden of rPTVs in childhood, persistent and late diagnosed ADHD

Whole exome-sequencing (WES) data were available for a subset of the iPSYCH samples. It has previously been shown that WES of DNA from dried blood spots results in high quality data^64^. DNA was extracted from dried blood spot samples of the study subjects and whole genome amplified in triplicates^50,51^, the coding regions of the genome were extracted using the llumina Nextera capture kit and sequencing was performed in multiple waves (Pilot 1, Wave 1, Wave 2 and Wave 3) using the Illumina HiSeq platform at the Genomics Platform of the Broad Institute.

Part of the data (Pilot 1, Wave 1, Wave 2) were also included in the recent study by Satterstrom et al.^35^, which looked at the overall burden of rPTVs in ADHD, and the same quality control procedure was used in this study, including all data (Pilot 1, Wave 1, Wave 2 and Wave 3). In short, the raw sequencing data were aligned to the reference genome Hg19 using the BWA^65^. Calling of genotypes was done using the best practice recommended by the Genome Analysis Toolkit^66^ (GATK) v.3.4. Most QC steps were performed using Hail 0.1 (Hail Team. Hail 0.2. https://github.com/hail-is/hail). All variants annotated to ACMG^67^ genes were removed due to Danish legislation. Samples were removed if they lacked complete phenotype information, sex inconsistencies of the imputed sex with the reported sex, if they were duplicates or genetic outliers identified by principal component analysis, if they had an estimated level of contamination >5% or if or if they had an estimated level of chimeric reads >5%. The number of individuals after QC were: childhood ADHD, N=4,987; persistent ADHD, N=748; late-diagnosed ADHD, N=1,915; controls, N=8,649.

Only autosomal genotypes were included in our analyses. Genotypes were removed if they did not pass GATK variant quality score recalibration (VQSR) or had read depth <10 or >1,000. Homozygous alleles were removed following if they had reference calls with genotype quality <25, homozygous alternate alleles with PL(HomRef) (i.e. the phred-scaled likelihood of being homozygous reference) less than 25 or <90% reads supporting alternate allele. Heterozygote alleles were removed if they had PL(HomRef) <25 or <25% reads supporting the alternate allele, less than <90% informative reads (i.e. number of reads supporting the reference allele plus number of reads supporting the alternate allele less than 90% of the read depth), or a probability of the allele balance calculated from a binomial distribution centered on 0.5 less than 1×10^−9^. After the application of these basic genotype filters, variants with a call rate <90% were removed, then samples with a call rate <95% were removed and then removal of variants with a call rate <95%. Between the sample call rate filter and the final variant call rate filter, one of each pair of related samples was removed, defining relatedness as individuals with a pair-wise pi-hat value ≥0.2. After QC, the number of individuals were: childhood ADHD, N=4,987; Persistent ADHD, N=748; Late-diagnosed ADHD, N=1,915; controls, N=8,649.

Following QC variants were annotated using SnpEff ^68^ version 4.3t with the command line ‘SnpEff GRCh37.75 input.vcf.gz’. The variants were also annotated with gnomAD^69^ exomes r2.1.1 database using SnpSift^68^ version 4.3t. Variants were only included if they were located in consensus high-confident coding regions with high read depth in both iPSYCH data and gnomAD dataset (80% of the samples in both datasets had at least 10× sequencing coverage in the region). Variants were defined as rPTVs if they were annotated as having large effects on gene function (nonsense variant, frameshift, splice site) and being rare in the sample defined as having an allele count no greater than 5 across the combination of our iPSYCH (n=16,299) and non-Finnish Europeans in the nonpsychiatric gnomAD exome database (n=44,779).

The burden of rPTVs and rSYNs in the three ADHD subgroups and controls were tested in two gene sets (1) Highly constrained genes (N=3,488), defined as genes being highly intolerant to loss of function mutations having a pLI score > 0.9^36^ (2) *de novo* highly constrained genes (N=241), defined as highly constrained genes which overlap with another set of genes (N=285) previously found to be enriched with *de novo* mutations in individuals with neurodevelopmental disorders^37^. For comparison we also tested a set of 9,662 evolutionary low constrained genes, i.e. having a pLI score < 0.1. Testing for enrichment in rPTVs and rare synonymous variants in the three groups compared to controls was done using multiple logistic regression with all groups included in the same regression model. We corrected the analyses using covariates: birth year, sex, first ten principal components from ancestry principal component analysis, number rSYN, percentage of target with coverage > 20x, mean read depth at sites within the exome target passing VQSR, total number of variants, sequencing wave. Testing for enrichment of rPTVs in ADHD subgroups compared to controls was corrected for nine tests (three groups x three gene sets, i.e. new P-value threshold=0.02), and tests for difference between groups were corrected for nine pair-wises comparisons (three gene-sets x three pair-wise comparisons for each set).

## Acknowledgements

The iPSYCH team was supported by grants from the Lundbeck Foundation (R102-A9118, R155-2014-1724 and R248-2017-2003), the EU FP7 Program (Grant No. 602805, “Aggressotype”) and H2020 Program (Grant No. 667302, “CoCA”), NIMH (1U01MH109514-01 to ADB) and the universities and university hospitals of Aarhus and Copenhagen. The Danish National Biobank resource was supported by the Novo Nordisk Foundation. High-performance computer capacity for handling and statistical analysis of iPSYCH data on the GenomeDK HPC facility was provided by the Center for Genomics and Personalized Medicine and the Centre for Integrative Sequencing, iSEQ, Aarhus University, Denmark (grant to ADB). M.S.A. is a recipient of a Juan de la Cierva Incorporación contract from the Ministry of Science, Innovation and Universities, Spain (IJC2018-035346-I). The research leading to these results has received funding from the Instituto de Salud Carlos III (PI17/00289, PI18/01788, P19/01224 and PI20/00041) and from the Agència de Gestió d’Ajuts Universitaris i de Recerca-AGAUR, Generalitat de Catalunya (2017SGR1461) and cofinanced by the European Regional Development Fund (ERDF). The Norwegian Mother, Father and Child Cohort Study is supported by the Norwegian Ministry of Health and Care Services and the Ministry of Education and Research. We are grateful to all the participating families in Norway who take part in this on-going cohort study. L.V.R. is a recipient of a pre-doctoral fellowship from the Instituto de Salud Carlos III, Spain (FI18/00285). M.R. was a recipient of a Miguel de Servet contract from the Instituto de Salud Carlos III, Spain (CP09/00119 and CPII15/00023).

## Conflicts of interest

D.D has received speaker fee from Takeda; J.A.R.Q was on the speaker’s bureau and/or acted as consultant for Janssen-Cilag, Novartis, Shire, Takeda, Bial, Shionogi, Sincrolab, Novartis, BMS, Medice, Rubiand Raffo in the last 3 years. He also received travel awards (air tickets + hotel) for taking part in psychiatric meetings from Janssen-Cilag, RubiShire, Takeda, Shionogi, Bial and Medice. The Department of Psychiatry chaired by him received unrestricted educational and research support from the following companies in the last 3 years: Janssen-Cilag, Shire, Oryzon, Roche, Psious, and Rubió.

## Author contributions

This section will be filled during the revision process

## Data availability

Summary statistics from GWAS of childhood, persistent and late-diagnosed ADHD is available at the iPSYCH website (https://ipsych.dk/en/research/downloads/)

## Correspondence and requests for materials should be addressed to

Ditte Demontis

## Notes

### Author Declarations

The Danish Scientific Ethics Committee, the Danish Health Data Authority, the Danish data protection agency, the Danish Neonatal Screening Biobank steering committee and Statistics Denmark, approved this study. The requirement of multiple permissions is in keeping with the ethical framework and the Danish legislation protecting the use of these samples.

