## Supplementary Figures for "Differences in the genetic architecture of common and rare variants in childhood, persistent and late-diagnosed attention deficit hyperactivity disorder"

**Supplementary Figure 1. Manhattan plots from GWAS of ADHD subgroups**

Results from logistic regression corrected for sex and ancestry principal components 1-10. Y-axes represent two-sided -log(P-values) and x-axes represent location on autosomal chromosomes. The red horizontal line represents the threshold for genome-wide significant association ($P = 5x10^{-8}$) **(A)** GWAS of childhood ADHD (14,878 cases; 38,303 controls) **(B)** GWAS of late diagnosed ADHD (6,961 cases; 38,303 controls) **(C)** GWAS of late diagnosed ADHD (1,473 cases; 38,303 controls).

**A.**

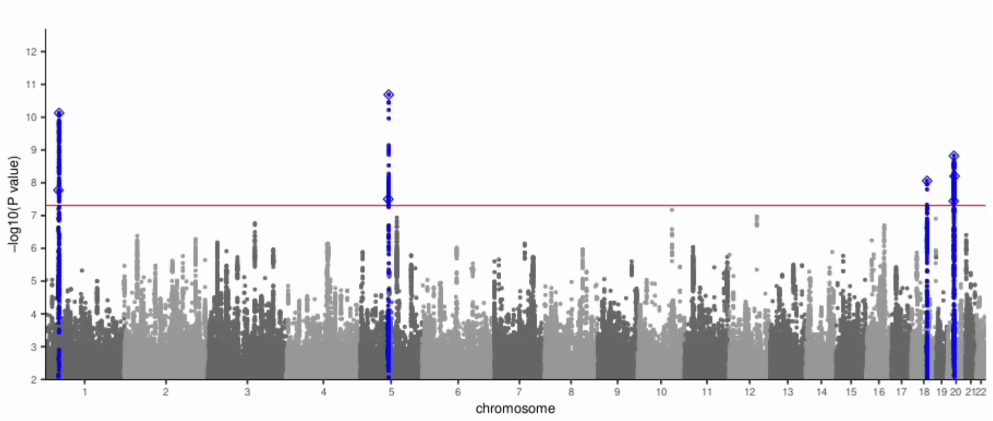

**B.**

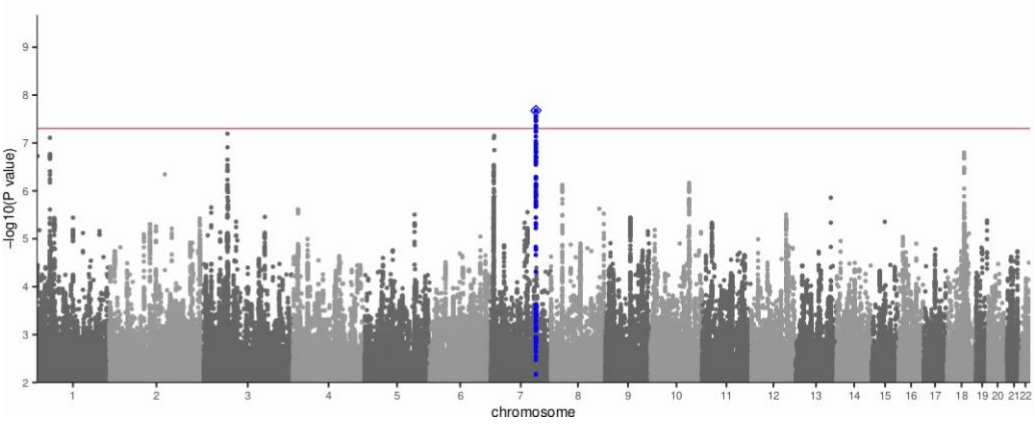

**C.**

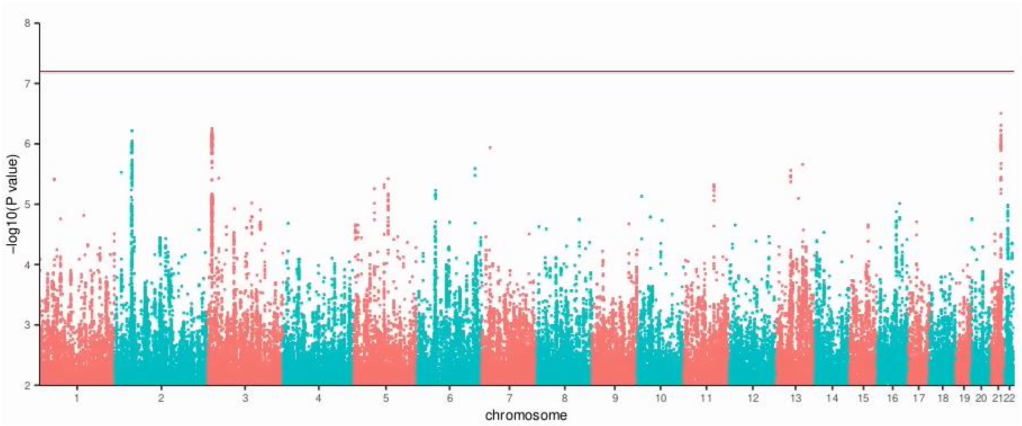

8
7
6
5
4
3
2
-log10(P value)
chromosome

**Supplementary Figure 2. Regional association plots of genome-wide significant loci**

Regional association plots of the local association results from the GWAS of ADHD sub-groups **(A-D)** the four genome-wide significant loci identified in the GWAS of childhood ADHD (14,878 cases; 38,303 controls) **(E)** the genome-wide significant locus identified in the GWAS of late-diagnosed ADHD. The y-axis represents –log(P-values) of variant association; the P-values are two-sided from logistic regression corrected using relevant covariates. Location and orientation of the genes in the regions are indicated on the X-axis, LD estimates of surrounding SNPs with the index SNP ($r^2$ values estimated based on 1KGP3) is indicated by colour (colour bar in upper left corner indicates $r^2$ values). Additionally, the local estimation of recombination rate is indicated in blue (legend on vertical axis at right).

**A.**

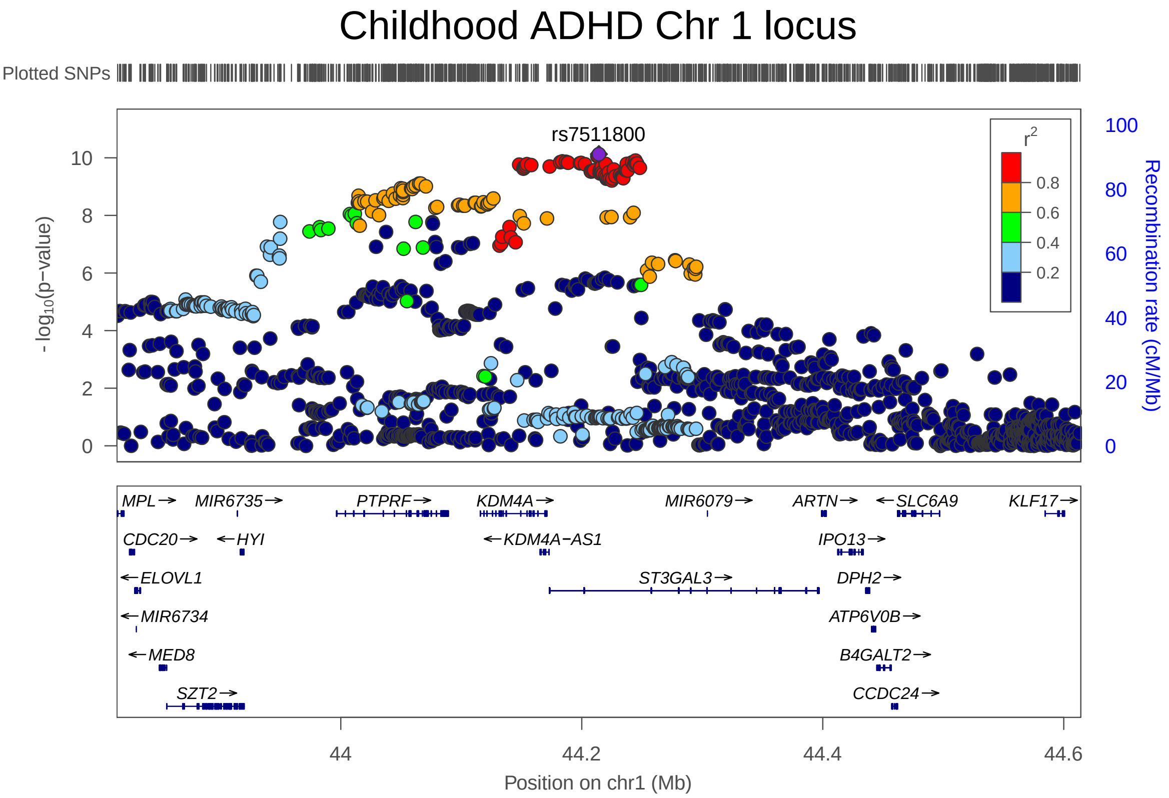

**B.**

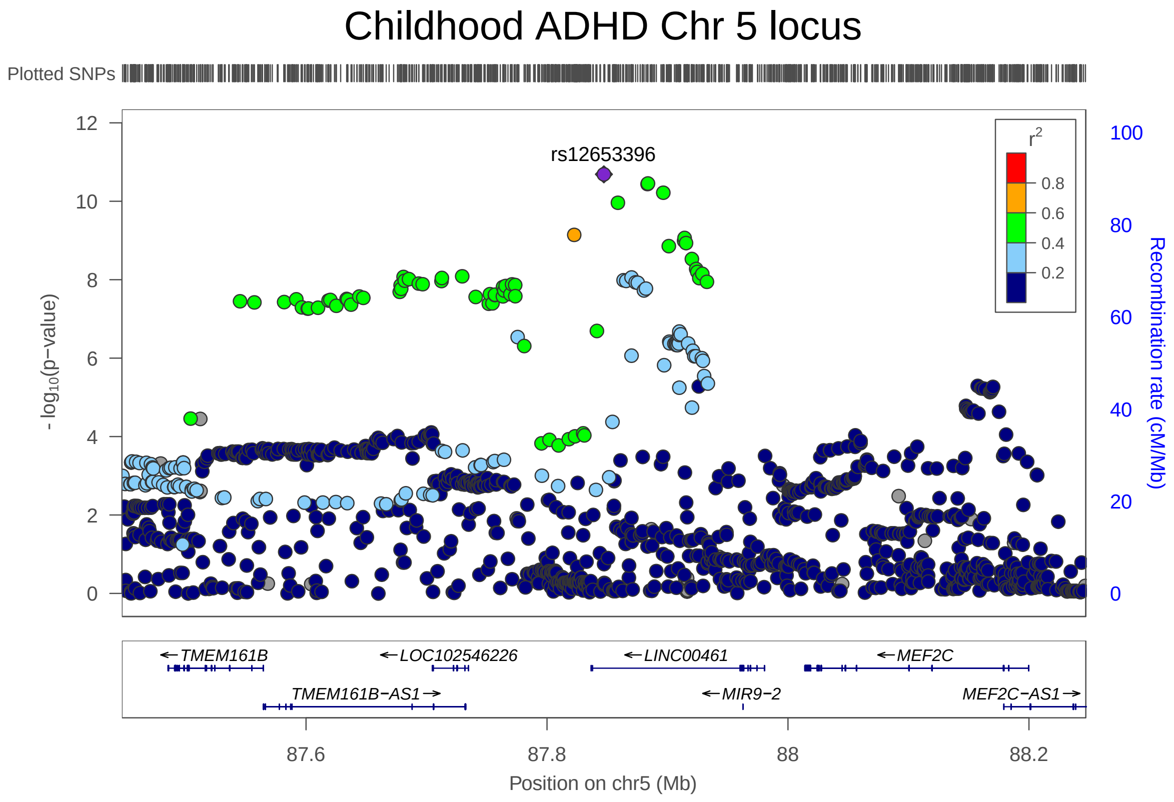

Childhood ADHD Chr 5 locus
Plotted SNPs
rs12653396
r2
0.8
0.6
0.4
0.2
- log10(p-value)
Recombination rate (cM/Mb)
TMEM161B
LOC102546226
LINC00461
MEF2C
TMEM161B-AS1
MIR9-2
MEF2C-AS1
87.6
87.8
88
88.2
Position on chr5 (Mb)

C.

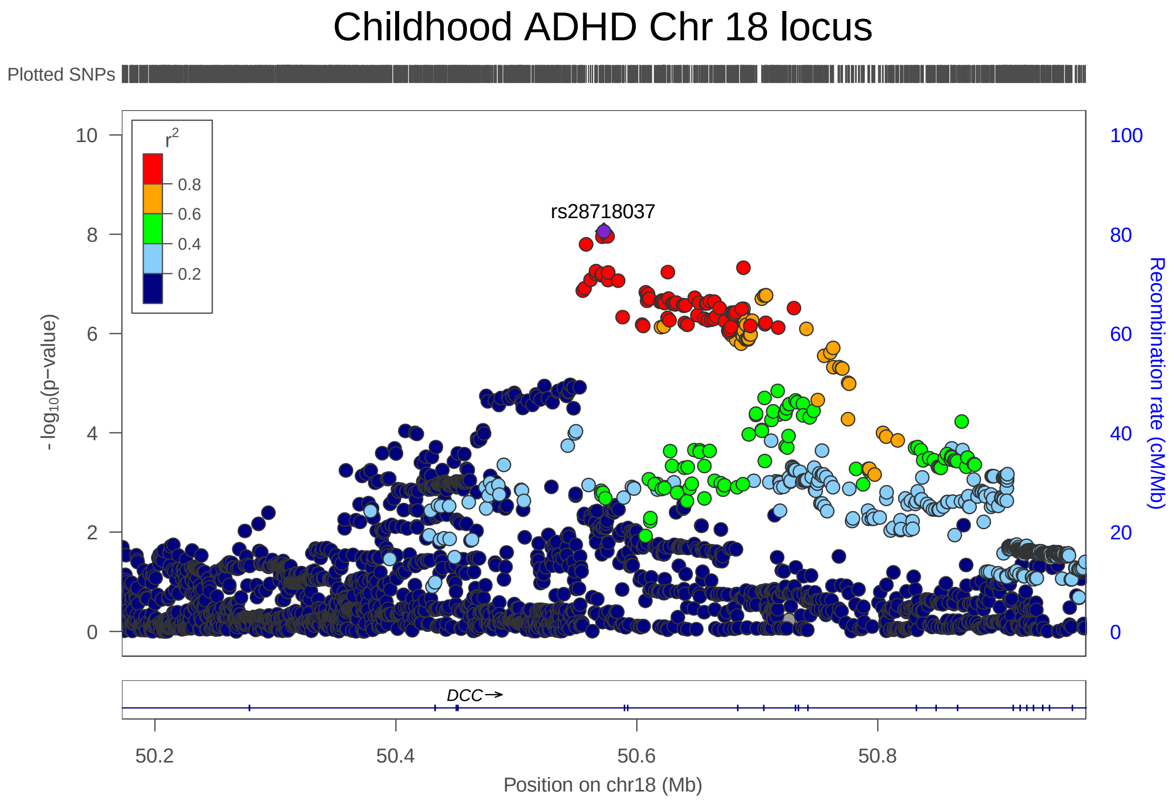

Childhood ADHD Chr 18 locus
Plotted SNPs
rs28718037
r2
0.8
0.6
0.4
0.2
- log10(p-value)
Recombination rate (cM/Mb)
DCC
50.2
50.4
50.6
50.8
Position on chr18 (Mb)

D.

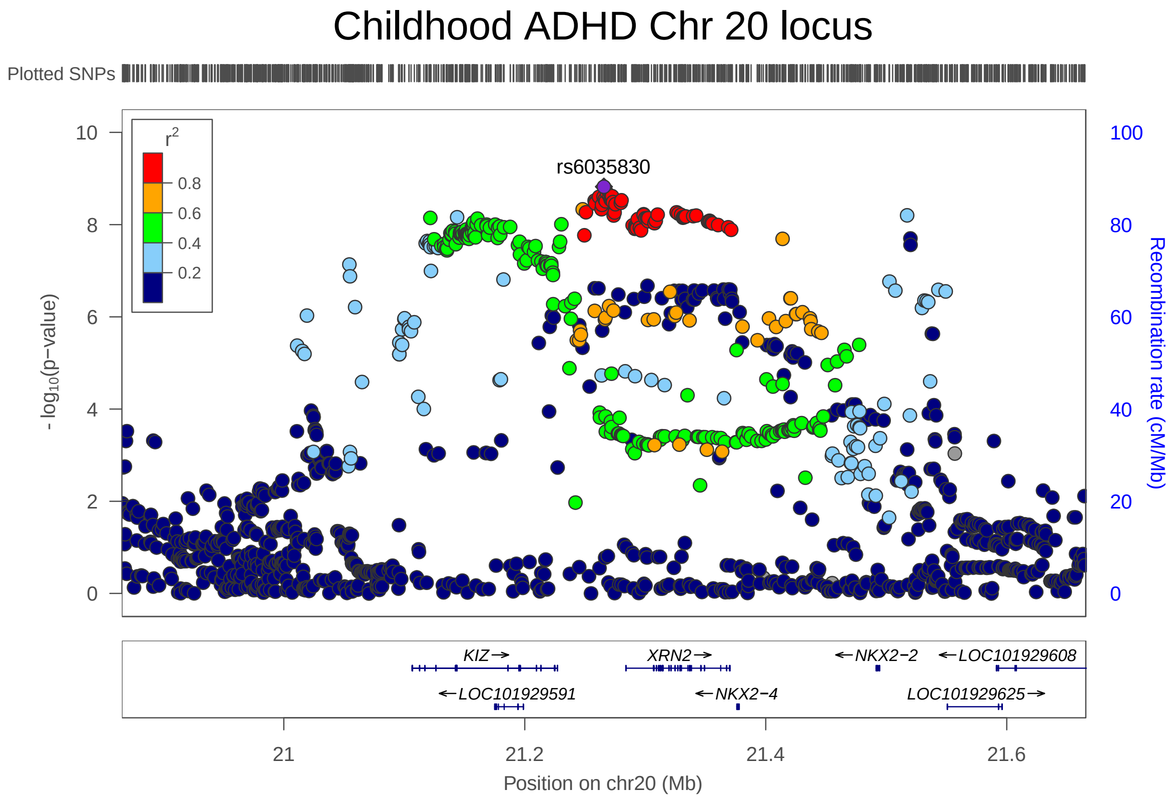
Childhood ADHD Chr 20 locus
Plotted SNPs
rs6035830
r2
0.8
0.6
0.4
0.2
- log10(p−value)
Recombination rate (cM/Mb)
KIZ→
XRN2→
←NKX2−2
←LOC101929608
←LOC101929591
←NKX2−4
LOC101929625→
21
21.2
21.4
21.6
Position on chr20 (Mb)

**E.**

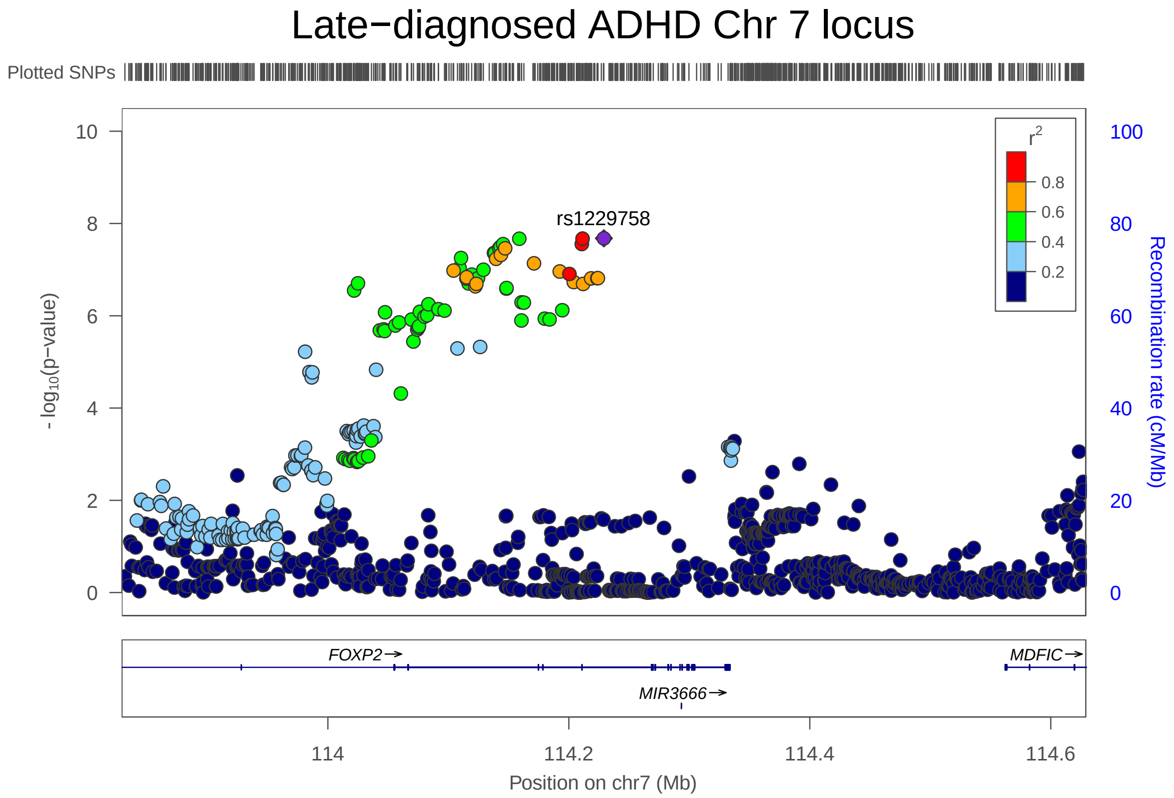
Late−diagnosed ADHD Chr 7 locus
Plotted SNPs
rs1229758
r2
0.8
0.6
0.4
0.2
- log10(p−value)
Recombination rate (cM/Mb)
FOXP2→
MDFIC→
MIR3666→
114
114.2
114.4
114.6
Position on chr7 (Mb)
